# Amyloid-PET pipeline choice influences classification of preclinical Alzheimer’s disease

**DOI:** 10.64898/2026.08.14.26360465

**Authors:** William Coath, Ariane Bollack, Catherine J Scott, Ashvini Keshavan, Ian B Malone, Heidi Murray-Smith, Pawel J Markiewicz, Kjell Erlandsson, Benjamin A Thomas, Frederik Barkhof, John C Dickson, Michael Schöll, the Insight 46 team, Jonathan M Schott, David M Cash

## Abstract

**BACKGROUND:** Quantitative amyloid-beta (Aβ)-PET is increasingly used in AD prevention trials. Although the Centiloid (CL) framework provides a common scale, variability persists across processing pipelines, including differences in template/native space, partial volume correction (PVC), and reference region. These choices may influence cut-points, and in turn positivity rates, as well as longitudinal accumulation rates. We examined cut-point estimates and inter-pipeline discordance in a community cohort where many are expected to have early Aβ deposition.

**METHODS:** We analysed [¹⁸F]florbetapir PET/MR data from predominantly cognitively unimpaired (∼95%) individuals aged ∼71 years at baseline (n=433) and at follow-up (n=328; ∼2.4-year interval) in Insight 46 (1946 British birth cohort). Centiloids were derived using the standard pipeline and ten in-house pipelines employing alternative reference regions and PVC in native space. Gaussian mixture modelling estimated cut-points with bootstrapped uncertainty. We assessed Aβ-discordance across pipelines as a function of standard CLs and examined follow-up CSF Aβ42/Aβ40 (n=120) and PET in individuals with discordant baseline classifications.

**RESULTS:** Baseline cut-points were 10–23 CL across pipelines, classifying 16–25% as Aβ-positive. Reliable accumulation cut-points were 3.5–6 CL/year, identifying 16– 22% as accumulators. Uncertainty varied across pipelines. At baseline, 18% were discordant across PET measures, predominantly between 11–35 standard CLs. The discordant group showed higher Aβ-PET accumulation and lower CSF Aβ42/Aβ40 than concordant negatives.

**CONCLUSIONS:** Disagreement between Aβ-PET methods was highest between 11–35 standard Centiloids and was frequently associated with accumulating Aβ. These findings highlight the importance of considering cut-point uncertainty and methodological influences when interpreting early-stage amyloidosis.

**Highlights:**

- Processing methods significantly influence amyloid PET Centiloid estimates
- Amyloid positivity thresholds and varied across commonly used PET methods
- PET discordance was greatest within an intermediate range between 11–35 Centiloids
- Future CSF and longitudinal PET suggest PET discordance frequently reflects early amyloidosis

## 1 Introduction

Recent trials of amyloid lowering therapies have demonstrated substantial reductions in amyloid-beta (Aβ) plaque and modest cognitive and functional benefits in individuals with early symptomatic Alzheimer’s disease (AD)[1,2]. As Aβ accumulates years before symptom onset, intervention during the preclinical stage may provide greater benefit^1^. Accordingly, secondary prevention trials of cognitively unimpaired individuals, such as AHEAD 3-45[3], increasingly rely on Aβ positron emission tomography (PET) or cerebrospinal fluid (CSF) Aβ42/Aβ40, typically following blood biomarker screening, to confirm evidence of Aβ pathology in preclinical disease. However, the reliable quantification of Aβ with PET is influenced by methodological factors that introduce variability across studies.

The Centiloid (CL) framework was developed to standardise Aβ PET measurements across tracers, studies, and pipelines by anchoring them to a common processing pipeline based on template space, fixed cortical target and whole-cerebellar reference regions, and the [11C]Pittsburgh Compound B (PiB) tracer[4]. This approach has been widely adopted as a common scale across tracers and studies[5]. However, reliance on template space can increase susceptibility to misregistration in individuals with atrophy, while image noise and scanner-specific characteristics may also influence Centiloid values[6,7]. Consequently, many groups employ alternative processing strategies, including native-space pipelines, partial volume correction (PVC), and different reference regions[7–10]. Although these methods aim to measure the same underlying biology, they can produce different Centiloid estimates and uncertainty, particularly for individuals within a 10–30 CL “intermediate zone”, where most Aβ positivity thresholds lie[5]. A recent meta-analysis of almost 49,227 Centiloids from 53 studies highlighted the variability in cut-points, that tend to fall within this intermediate zone, but a broad approach is unable to examine how differences in processing may contribute towards this heterogeneity[11].

Aβ positivity thresholds may be derived from visual reads, neuropathology, prediction of future accumulation, or statistical approaches such as Gaussian mixture modelling (GMM)[12–15]. Because these thresholds variably depend on study population, timing and processing methodology, cut-points and their associated uncertainty vary substantially. Consequently, individuals near the threshold may be classified differently across pipelines despite measuring the same underlying pathology. Determining whether such discordance reflects biological heterogeneity, measurement noise, or methodological artefacts requires longitudinal and multimodal evaluation.

Longitudinal PET can distinguish true Aβ accumulation from measurement variability in individuals near a positivity threshold. Combined with CSF Aβ42/Aβ40, which often becomes abnormal earlier than PET, it can help determine whether discordant PET classifications reflect emerging pathology or methodological artefacts[16]. This is particularly informative in individuals whose Aβ status differs across processing methods.

Insight 46, the neuroimaging sub-study of the MRC National Survey of Health and Development (the 1946 British birth cohort), provides an ideal setting to address these questions[17,18]. As a population-derived, nationally representative cohort with a narrow age range, it minimises the confounding effects of age heterogeneity common in ageing studies[6]. Participants underwent longitudinal Aβ PET imaging between ages 69 and 73 years on a single scanner, together with CSF sampling, providing a unique opportunity to evaluate methodological influences on Aβ quantification and the implications of discordant PET classifications.

In this study, we apply this framework to compare baseline and longitudinal Aβ PET across multiple processing pipelines in Insight 46. We expect that the cohort comprises a mixture of individuals with and without Aβ pathology that can be separated using GMM. Our aims are to: (i) characterise the influence of PET processing methodology on Centiloid cut-point estimates and their uncertainty for defining Aβ positivity and reliable Aβ accumulation; (ii) determine how discordance in Aβ status across pipelines varies according to Centiloids derived from the standard Centiloid method; and (iii) investigate the CSF Aβ42/Aβ40 and longitudinal Aβ PET profiles of individuals with discordant baseline Aβ PET classifications.

## 2 Methods

### 2.1 Participants

Participants were enrolled into Insight 46, with data collected in two phases in London (UK) and study protocols for phases 1 and 2 were described in detail previously[17,18].

### 2.2 Image acquisition

PET and MRI data were acquired on a single 3T Siemens Biograph mMR PET/MR scanner at both timepoints. PET data were acquired for 60 mins following injection of ∼370 MBq [^18^F]florbetapir (AV45). During the PET acquisition, 3D T1-weighted magnetization-prepared rapid acquisition gradient echo (MPRAGE, 1.1 mm isotropic resolution) and 3D T2-weighted (1.1 mm isotropic resolution) images were acquired. Subject specific pseudo-CT (pCT) images were synthesised through matching of T1 and T2-weighted MRIs to a CT database for attenuation correction (AC)[19], a method which has been thoroughly validated across multiple datasets to produce robust and accurate AC maps that are comparable to CT-based AC[20]. List mode PET data from 50 to 60 minutes post-injection were reconstructed using pCT AC with an ordered subsets expectation maximisation (OSEM) algorithm with 4 iterations and 14 subsets (voxel size 2 mm isotropic), implemented with open source NiftyPET software[21].

### 2.3 Image processing

PET images were processed to derive standard Centiloid template-based SUVRs for reference, and separately with ten non-standard pipelines using a native space approach.

#### 2.3.1 Standard approach

The Insight 46 dataset was processed using a local version of the standard Centiloid statistical parametric mapping (SPM) pipeline described in Klunk et al.^4^ and validated previously through replication of the “Level 1” analysis using the publicly available Standard PiB dataset (R^2^ = 0.9994)[8]. The standard pipeline uses WC reference and cortical target regions in SPM’s MNI-152 space to derive SUVRs; we refer to this measure as ‘STDWC’.

#### 2.3.2 Non-standard native space approach

T1-weighted MRI scans were parcellated using the Geodesic Information Flow (GIF) algorithm[22]. An additional processing step to exclude remaining non-brain tissue was performed using a whole brain mask generated with multi-atlas propagation segmentation (MAPS) which was visually reviewed and edited where necessary by a trained rater[23]. The T1-weighted MRI was rigidly registered to the PET image using a NiftyReg[24]. We then produced SUVR images both with and without PVC. For non-PVC images, the GIF labels were resampled to PET space. For PVC images, the PET was instead resampled to T1-space and iterative Yang PVC was performed with a 6.8 mm isotropic Gaussian kernel using the PETPVC toolbox[9,25]. SUVR images were then created using five reference regions: i) cerebellar grey matter (CGM), cleaned with a grey matter tissue probability mask threshold at 0.9; ii) whole cerebellum (WC); iii) subcortical white matter (WM), eroded by one voxel in the final image space (i.e. larger PET voxel erosion for non-PVC images) to reduce partial volume effects; iv) the pons, also eroded with the same approach; and v) a composite reference region of all (Figure S1). Mean SUVR values were then extracted from a large cortical summary GIF target region including frontal, cingulate, parietal, and lateral temporal cortical areas^26^. These measures will be referred to as ‘REFERENCE{_PVC}’ (e.g. WC for whole cerebellum reference without or WC_PVC with PVC).

#### 2.3.3 Centiloid conversion

The resulting SUVRs from all pipelines were converted to Centiloids using a previously described approach and extended here to incorporate additional reference regions (CGM, Pons and composite) and updated equations for NiftyPET reconstructed images[8]. Centiloid equations for all pipelines are presented in Table S1.

### 2.4 Non-imaging measures

Additional participant data included *APOE* ε4 genotyping, cognitive and clinical assessments[17,18]. CSF samples were collected via lumbar puncture on a subsample of participants at phase 2 of Insight 46 (contemporaneous with the follow-up PET timepoint). CSF Aβ42 and Aβ40 were measured using Lumipulse assays (Fujirebio) as previously described[26].

### 2.5 Statistical analysis

#### 2.5.1 Defining Aβ positivity and reliable accumulation

First, to assess the similarity between different PET measures, pairwise non-parametric Spearman’s rank correlation coefficients were computed with significance level set at *p* < 0.05 (uncorrected for multiple comparisons). For cut-point definition, GMM was implemented in R with the *‘mclust’* package[27] to fit the overall distribution of baseline Centiloid values (for positivity) and annual rate of change (ARC in CL/year for reliable accumulation) from each measure. Models with one, two and three Gaussians were compared with Bayesian information criterion (BIC) to determine which model provided the best fit. As the hypothesised large ‘Aβ negative’ Gaussian should be robust in a mainly cognitively unimpaired sample, the cut-point value was defined as the 99^th^ percentile (z-score = 2.326) of this lower distribution. To assess the stability of cut-points from different measures, bootstrapping was performed in *mclust* to replicate the GMM and calculate 5000 estimates of the cut-point value for each pipeline. The 95% confidence intervals (CIs) of this cut-point distribution were calculated and applied to the original sample to generate lower-and upper-percent positive rate estimates. A narrow CI range suggests the cut-point is robust to sampling variation with reliable separation of negative and positive cases, whereas a wide CI range indicates a less robust cut-point.

Agreement between pairs of measures in assessment of binary status (e.g. Aβ status at baseline or accumulation status) was assessed with Cohen’s unweighted kappa (κ) coefficient[28] implemented with the *kappa2* function from the ‘*irr*’ R package. A κ score of 1 indicates perfect agreement between measures whereas 0 corresponds to chance level agreement, negative values are possible if agreement is worse than chance.

#### 2.5.2 Defining PET discordance across pipelines

To characterise Aβ PET status discordance, all PET scans at both timepoints were categorised into one of three groups based on assigned Aβ positivity status across all eleven measures (including the standard approach): (1) concordant negative with all measures (ConcN), (2) Discordant across measures (Disc; positive on at least one measure but not all), and (3) concordant positive with all measures (ConcP). Chi-squared tests were used to test for significant differences between concordance categories by APOE ε4 carriership and sex (significance level set to *p* < .05).

#### 2.5.3 Baseline PET discordance by standard Centiloid value

We investigated the probability of a scan being assigned to each concordance category (ConcN, Disc or ConcP) as a function of the standard Centiloid (CL_STDWC)_ value of the scan. A multinomial logistic regression model was used to estimate the probability of a scan belonging to each concordance category as a function of CL_STDWC_ (*multinom* in the *nnet* R package, formula: *concordance_category ∼ CL_STDWC_*). Centiloid values where the category probability curves cross (i.e. the most likely category changes) were identified and may be informative for understanding how the uncertainty in Aβ status changes across different levels of pathology. We used the CL_STDWC_ approach as a reference measure to investigate discordance between measures not because it is expected to be the optimal method but because it is widely used and reported so provides useful contextual information.

#### 2.5.3 Transitions between PET concordance categories over time and accumulation rates

We investigated the stability of concordance categories over the follow-up period and report how many individuals advanced (ConcN->Disc, Disc->ConcP, ConcN->ConcP; increasing evidence of Aβ) or reverted categories (Disc->ConcN, ConcP->Disc, ConcP->ConcN; decreasing evidence of Aβ). ARC in CL/year were compared across the three baseline concordance groups for each measurement approach using ANOVA and post-hoc tests with Benjamini-Hochberg false discovery rate (FDR) correction, with Welch’s ANOVA and Games-Howell post-hoc tests used in the case of unequal variances (Levene’s test *p* < 0.05). The proportion of reliable accumulators was compared between each of the baseline concordance categories using Fisher’s exact tests (FDR corrected).

#### 2.5.3 PET discordance and CSF Aβ42/Aβ40

We then investigated CSF Aβ42/Aβ40 ratio at follow-up by concordance category at both timepoints. Individual PET positivity status across measures was visualised in individuals that were in the discordant category at either timepoint. CSF Aβ42/Aβ40 was compared between baseline concordance groups using ANOVA. Although CSF is treated as a continuous measure in this analysis, for reference purposes, we show published FDA positive (ratio ≤0.058) and negative (ratio ≥ 0.073) thresholds, with likely positive cases in the 0.059–0.072 range.

## 3 Results

Participant characteristics for baseline and longitudinal samples are presented in Table 1.The longitudinal sub-sample has similar characteristics to the full baseline sample. While cases of dementia were very rare across timepoints (n = 2, 0.6%), there was a numerical increase in cases of MCI from baseline (n = 5, 1.5%) to follow-up (n = 14, 4.3%).

**Table 1.**
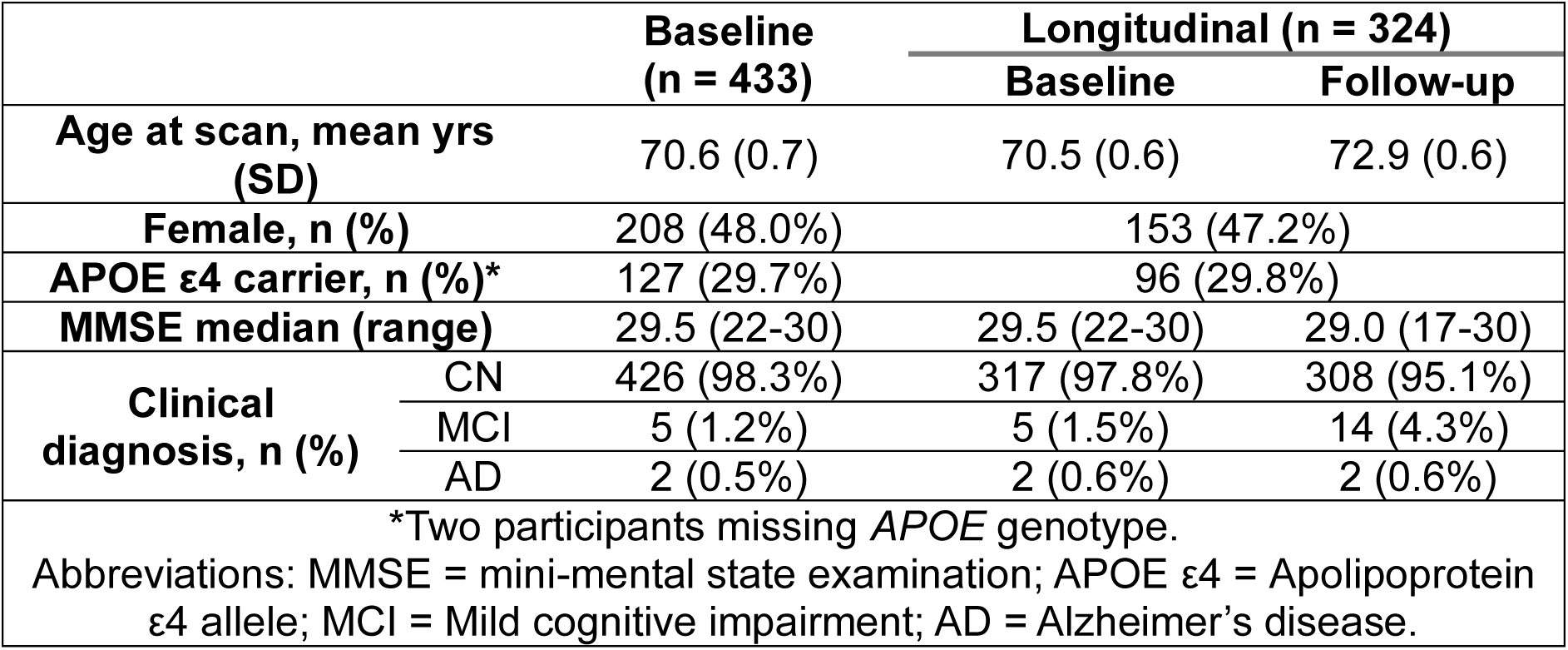
Participant characteristics at baseline and in the longitudinal group

### 3.1 Similarity of measures

At baseline (Figure S2A), CL_STDWC_ was most strongly correlated with CL_WC_ (ρ = 0.95) and most weakly correlated with CL_WM_PVC_ (ρ = 0.44). As expected, associations tended to be the strongest between SUVRs when either both were based on the cerebellum (ρ = 0.66–0.95), or both were based on white matter reference regions (ρ = 0.67-94). The weakest correlations were observed between CL_CGM_ and white matter-referenced CLs (ρ = 0.33–0.46). For ARC (Figure S2B), the strongest corelation with CL_STDWC_ was also the CL_WC_ approach (ρ = 0.97) whereas correlations with CL_WM_ and CL_WM_PVC_ were statistically non-significant (*p* > .05, ρ < 0.08). Reference region had a large impact on correlations between non-standard measures, with very weak or negative (ρ < 0.07) correlations between CL_CGM_ and the following measures: CL_WM_, CL_WM_PVC_, CL_Pons_, CLPons_PVC, CL_Comp_PVC_.

### 3.2 Aβ positivity and reliable accumulation

#### 3.2.1 Aβ positivity

For each measure, the fit of GMMs using one to three Gaussian components with equal and unequal variances were compared with BIC (Table S2). In all cases, two Gaussians provided a substantially better fit than one Gaussian and unequal variances between distributions provided a better fit than equal variance. Most measures were best fit with a bi-modal unequal variance model. For CL_CGM_PVC_, CL_WC_PVC_ and CL_Pons_PVC_, the fit for the three Gaussian unequal variance model was marginally better than two Gaussians. However, the difference in BIC between two and three Gaussian models was substantially smaller than the difference observed between one and two Gaussian models. Therefore, to keep cut-points and other metrics comparable across measures, bi-modal unequal variance GMMs were fitted for all measures (see Figures S3 and S4). Centiloid cut-points were then defined at the 99th percentile of the lower distribution, falling between 10.0 [7.4, 13.5] CL (CL_Comp_) and 22.9 [19.0, 27.4] CL (CL_WM_) (Figure1A and numerically in Table S3). For all reference regions apart from the composite, PVC slightly lowered the cut-points (with overlapping 95% CI). WM and Pons referenced measures tended to have higher cut-points compared to measures containing the cerebellum in the reference. With PVC applied, the uncertainty (range between lower and upper 95% CI; Figure 1B) in cut-points was numerically lower for measures with cerebellum reference regions (CL_CGM_ = 10.3 vs CL_CGM_PVC_ = 6.1; CL_WC_ = 6.5 vs CL_WC_PVC_ = 4.9), very similar for WM and Pons (CL_WM_ = 8.4 vs CL_WM_PVC_ = 9.2; CLPons = 8.6 vs CL_Pons_PVC_ = 8.1) and higher for the Composite reference (CL_Comp_= 6.1 vs CL_Comp_PVC_ = 9.0).

**Figure 1.**
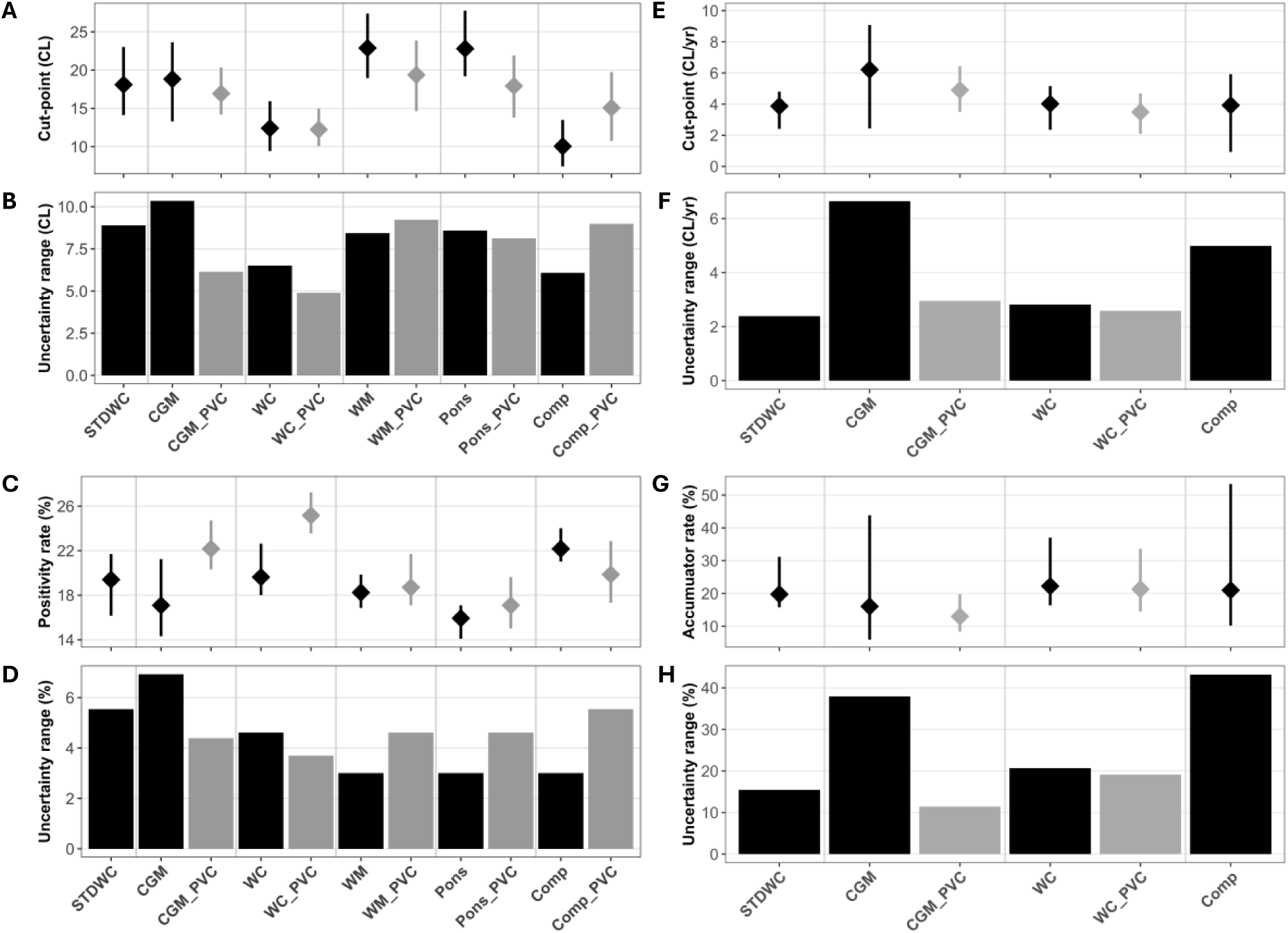
Comparison of cut-points and baseline positivity (A-D) and longitudinal accumulation (E-H) across Centiloid measures. Non-PVC measures are shown in black and PVC are in grey. **A**: Positivity cut-point estimates in Centiloids. **B**: Comparison range of uncertainties in positivity cut-point estimates (height of bars in panel B matches error bars in A). **C**: Positivity rate estimates (% of baseline sample rated positive). **D**: Range of uncertainty in positivity rate estimates. **E**: Reliable accumulation cut-point estimates in Centiloids per year. **F**: Range of uncertainty in reliable accumulation cut-points. **G**: Reliable accumulation rate estimates. **H**: Range of uncertainty in accumulation rate estimates. STDWC = standard whole cerebellum; CGM = cerebellar grey matter; WC = whole cerebellum; WM = white matter; Comp = Composite; PVC = partial volume correction.

When applying these cut-points to assign Aβ status, there was generally substantial pairwise agreement between all measures (all κ ≥ 0.66). Agreement with the CL_STDWC_ was highest for CLWC (κ = 0.95) and lowest for CL_WM_PVC_ (κ = 0.71). Agreement with and without PVC for a given reference region was very good (CGM = 0.83, WC = 0.83, WM = 0.92, Pons = 0.87, Comp = 0.86) and agreement tended to be lower between methods using different reference regions and PVC (Figure S5A). Positivity rate estimates (%) and associated uncertainty are compared in Figure 1C and D and numerically in Table S3, these ranged between 15.9% (CL_Pons_) to 25.2% (CL_WC_PVC_). For CGM and WC reference regions, PVC tended to increase the positivity rate (CL_CGM_ = 17.1% [14.3, 21.2] vs CL_CGM_PVC_ = 22.2% [20.3, 24.7]; CL_WC_ = 19.6% [18.0, 22.6] vs CL_WC_PVC_ = 25.2% [23.6, 27.3]; Figure 1C). For WM and Pons, positivity rates were similar with PVC (CL_WM_ = 18.2% [16.9, 19.9] vs CL_WM_PVC_ = 18.7% [17.1, 21.7]; CLPons = 15.9% [14.1, 17.1] vs CL_Pons_PVC_ = 17.1% [15.0, 19.6]). For the composite reference, the positivity rate was lower with PVC (CLComp = 22.2% [21.0, 24.0] vs CL_Comp_PVC_ = 19.9% [17.3, 22.9]). The highest level of uncertainty in the positivity rates (Figure 1D) was observed for CLCGM (95% CI: 13.9-21.2%, range = 7.3%), while the lowest level of uncertainty was found for CL_Pons_ (95% CI: 14.3-17.1%, range = 2.8%). With PVC applied, the uncertainty was lower in measures using cerebellar reference regions compared to the non-PVC measure (CL_CGM_ = 6.9% vs CL_CGM_PVC_ = 4.4%; CL_WC_ = 4.6% vs CL_WC_PVC_ = 3.7%) and higher when using the other reference regions (CLWM = 3.0% vs CL_WM_PVC_ = 4.6%; CLPons = 3.0% vs CL_Pons_PVC_ = 4.6%; CL_Comp_ = 3.0% vs CL_Comp_PVC_ = 5.5%).

#### 3.2.3 Reliable Aβ accumulation

For ARC, CGM and WC referenced measures were best fit by two components with unequal variance terms. The CL_Comp_ measure was also best fit by two components with similar BIC for equal and unequal variance (see Table S4). Unlike the cross-sectional analysis, there were no instances of a three-component model providing the best fit. CL_WM_, CL_WM_PVC_, CL_Pons_, CL_Pons_PVC_ and CL_Comp_PVC_ measures were best fit with unimodal models and therefore could not be used to define a reliable accumulation threshold with a bi-modal unequal variance GMM. These measures which favoured the single Gaussian model were excluded from the longitudinal analysis. GMM fits for ARC with all measures are presented in Figures S6 and S7. For CLSTDWC, CL_CGM_, CL_CGM_PVC_, CL_WC_, CL_WC_PVC_ and CL_Comp_ measures, bimodal GMMs were fit to ARC, reliable accumulation cut-points were placed at the 99th percentile of the lower component. Figure 1E and F show accumulation cut-points (CL/year) with associated uncertainty (95% CI from bootstrapping) for each measure. Cut-point estimates fell between 3.5 [2.1, 4.7] CL/year (CL_WC_PVC_) and 6.2 [2.4, 9.1] CL/year (CL_CGM_). Uncertainty in cut-points was highest for CL_CGM_ and CL_Comp_ measures. All reliable accumulation cutpoints are presented numerically in Table S3.

Next, accumulator status was assigned to all individuals with each measure if their ARC was above the reliable accumulation cut-point for that measure. Pairwise agreement (kappa) in accumulator status was generally moderate (Figure S5B). Agreement with the CL_STDWC_ measure was highest with CL_WC_ (κ = 0.87) and lowest with CL_Comp_ (κ = 0.56). Accumulator rate estimates (%) and the uncertainty in these estimates (rates from application of lower and upper 95% CI cut-points) are presented in Figure 1G and H. The prevalence of accumulators ranged between 13.0% [8.3, 19.8] with CL_CGM_PVC_, to 22.2% [16.4, 37.0] with CL_WC_. The uncertainty range in the accumulator rates was highest for CL_Comp_ (95% CI: 10.2-53.4%, range = 43.1%), while the estimate with the lowest uncertainty was CLCGM_PVC (95% CI: 8.3-19.8%, range = 11.5%). Accumulator rates are presented numerically in Table S3.

### 3.3 PET discordance at baseline

#### 3.3.1 PET discordance at baseline

Scans were then classified based on the concordance of assigned PET status across measures. At baseline (n = 433), individuals were classified as ConcN (n = 306, 70.7%), Disc (n = 76, 17.6%) or ConcP (n = 51, 11.8%). The proportion of APOE ε4 carriers was significantly different between categories (Chi-square, n = 431, *χ*^2^ = 58.54, *p* < 0.001). Post-hoc tests showed that the proportion of carriers significantly increased from ConcN to Disc (19.4% to 46.1%; n = 380, *χ*^2^ = 21.78, *p* < 0.001) and from Disc to ConcP (46.1% to 66.7%; n = 127, *χ*^2^ = 4.43, *p* = 0.035). The proportion of females across concordance categories was similar with no significant sex difference observed (ConcN = 48.0%, Disc = 46.1%, ConcP = 51.0%; n = 433, *χ*^2^ = 0.30, *p* = 0.862; see Figure S8).

#### 3.3.2 Baseline PET discordance by standard Centiloids

### Error! Reference source not found

A shows the distributions of CL_STDWC_ values in each concordance category at baseline. Probabilities from logistic regression indicate that a scan was most likely to be classified as discordant between 11–35 CL_STDWC_, with 41 (53.9%) of all discordant scans falling within this range (Figure 2B). Below 11 CLSTDWC, 296 (90.5%) of all scans were ConcN and 31 (9.5%) were Disc, with no ConcP cases. Between 11–35 CL_STDWC_, 10 (17.2%) were ConcN, 41 (70.7%) were Disc and 7 (12.1%) were ConcP. Above 35 CL_STDWC_, none were ConcN, 4 (8.3%) were Disc and 44 (91.7%) were ConcP. The range of ConcN values was -35– 17 CL_STDWC_ and the range of ConcP was 25–131 CLSTDWC . Discordance was observed between -32–51 CL_STDWC_ (below -32 CL_STDWC_ all scans were classified ConcN, and above 51 CL_STDWC_ all were ConcP). The heatmap presented in Figure 2C shows Aβ PET status from each individual measure ordered by CL_STDWC_, revealing which measures are responsible for discordance (rows that are not completely orange or grey) across the range of standard Centiloid values. Measures combining cerebellum reference and PVC, or composite reference classified most individuals in the 11–35 CLSTDWC range as positive (CL_WC_PVC_ = 74%, CL_CGM_PVC_ = 67%, CLComp = 64%), whereas non-cerebellar reference regions with PVC classified less than half as positive (CL_WM_PVC_ = 33%, CL_Pons_PVC_ = 34%). In the 0–11 CL_STDWC_ range (n = 78) measures with PVC or composite tended to classify more scans as positive (CLWC_PVC = 19%, CL_WM_PVC_ = 13%, CLComp_PVC = 13%, CL_Comp_ = 12%) and whereas cerebellum without PVC classified the least (CL_STDWC_ = 0%, CL_WC_ = 1%, CL_CGM_ = 3%). Positivity was rare below 0 CLSTDWC (n = 249), with most cases from measures with WM containing reference regions or with PVC applied (CL_WM_PVC_ = 2%, CL_WM_ = 2%, CL_Comp_PVC_ = 2%, CLCGM_PVC = 1%, CL_WC_PVC_ = 1%, CLPons, CLPons_PVC and CL_Comp <_ 1%) and none with CL_STDWC (_by definition), CL_CGM a_nd CL_WC m_easures.

**Figure 2.**
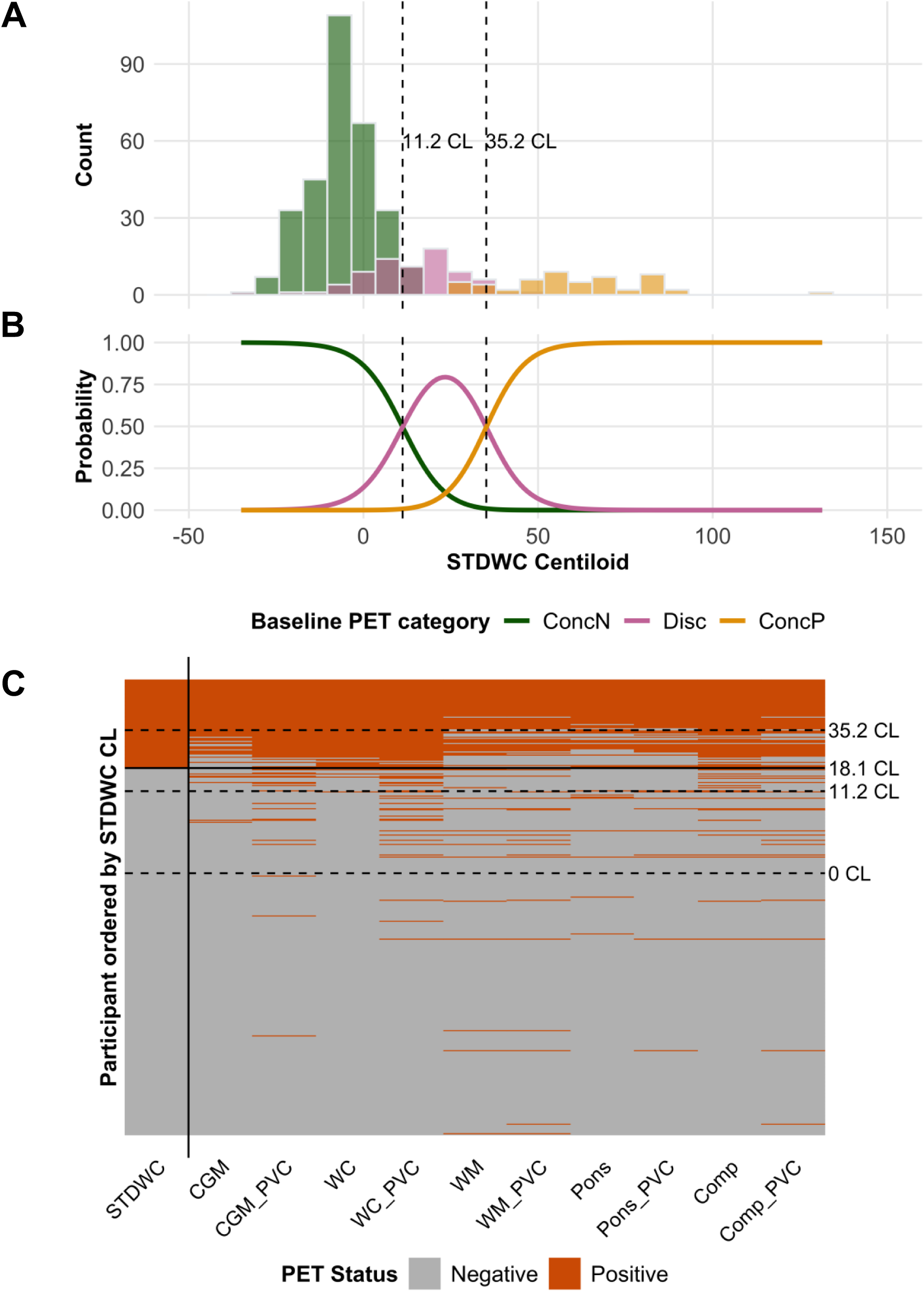
Baseline PET status discordance across measures (amyloid positivity in at least one measure but not all) as a function of Centiloid value derived from the standard method (STDWC). **A**: Histogram showing the distribution of each baseline PET concordance category across STDWC Centiloid values. **B**: Logistic probability curves for each category across STDWC Centiloid values. Dashed vertical lines on A and B are boundary values where probability of PET status discordance becomes most likely (11 – 35 CL). **C**: Heatmap showing individual amyloid positivity across measures (x) with participants ordered by STDWC (y) with solid line representing the STDWC GMM derived cut-point at 18 CL and dashed lines representing boundary values (11-35 CL from panels A and B) as well as 0 CL for reference.

### 3.4 Transitions between PET concordance categories over time and accumulation rates

For those who had scans at both timepoints (n = 324), at baseline 234 individuals (72.2%) were classified as ConcN, 57 (17.6%) were Disc and 33 (10.2%) were positive across all measures. The flow of individuals transitioning between concordance categories over the follow-up interval using the baseline pipeline specific positivity thresholds at both timepoints is presented in Figure 3A. No individuals transitioned directly between ConcN and ConcP in either direction. Of the ConcN individuals at baseline, 213 (91.0%) remained ConcN and 21 (9.0%) advanced to Disc at follow-up. For the 57 individuals classified as Disc at baseline, 14 (24.6%) advanced to ConcP and only three (5.3%) reverted to ConcN at follow-up, with 40 (70.2%) remaining Disc. Of the 33 individuals who were ConcP at baseline, only one reverted to Disc, while the others remained at ConcP.

**Figure 3.**
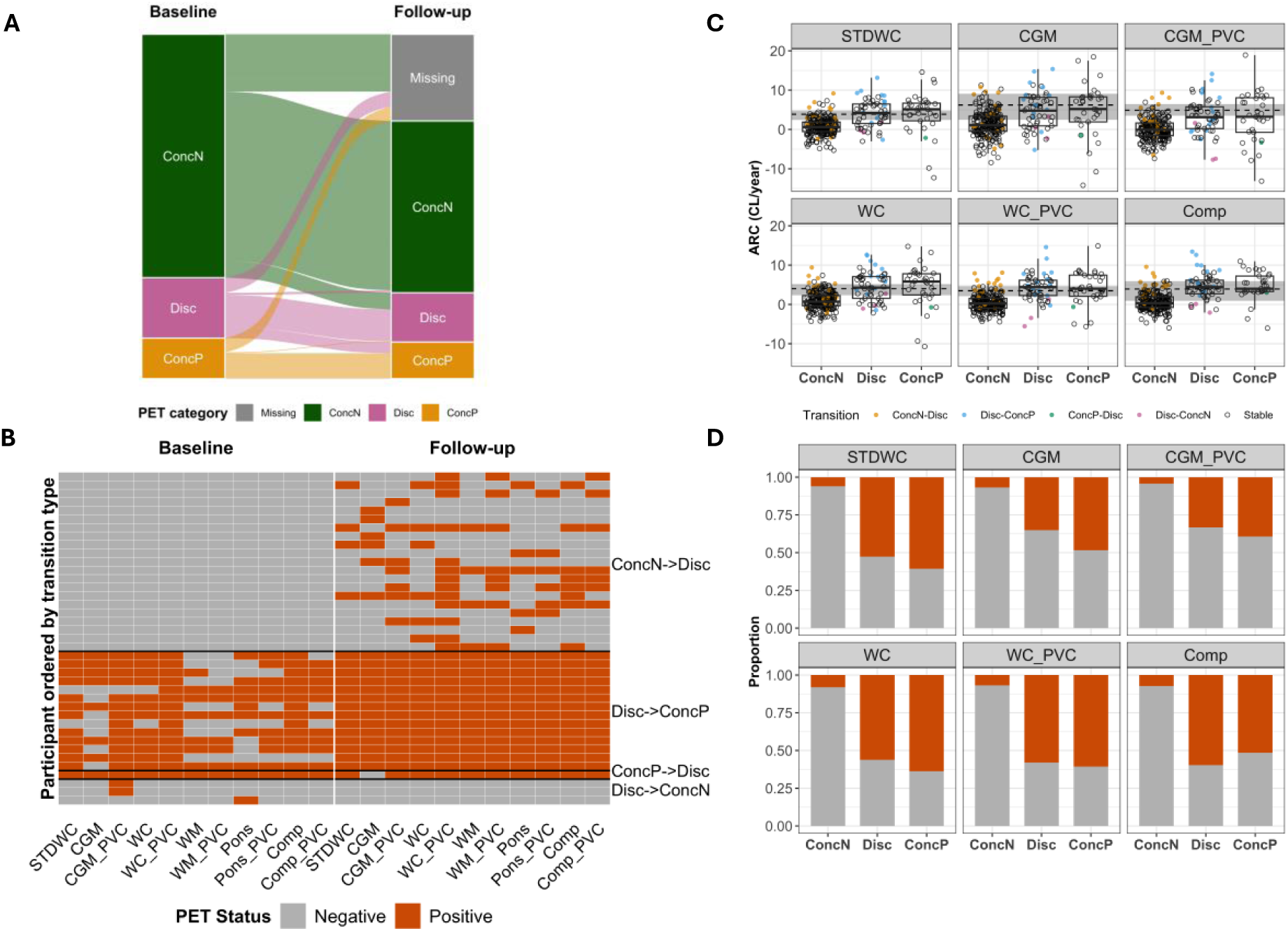
Discordance across PET pipelines over time. **A:** Transitions between PET concordance categories at baseline and follow-up. **B:** The colour of each cell represents PET status at baseline (left) or follow-up (right) across measures in individuals who either advanced (ConcN->Disc, Disc->ConcP) or reverted (ConcP->Disc, Disc-ConcN) categories over time. **C:** Annual rate of change (ARC) in Centiloids between timepoints for each measure by baseline PET concordance category. Gaussian mixture modelling reliable accumulation threshold is represented by the black dashed line with shaded region for 95% CI. Points are coloured by transition category. **D:** The proportion of reliable accumulators for each measure (above ARC cut-points in C) by baseline PET concordance.

Figure 3B shows the individual positivity across measures at baseline and follow-up in individuals who transitioned between categories. A transition from Disc to ConcP is suggestive of early detection in measures showing baseline positivity. The CL_WC_PVC m_easure classified all 14 individuals who advanced from Disc to ConcP as positive at baseline whereas CL_WM_PVC_ only classified 7/14 as positive at baseline. Reversion (Disc to ConcN or ConcP to Disc) based on a single measure is suggestive of a false positive/negative classification for that measure. The three individuals who reverted from Disc to ConcN were categorised as positive at baseline only by one of the measures: CL_CGM_PVC_ (n = 2) and CL_Pons_ (n = 1). The one individual who reverted from ConcP to Disc was negative at follow-up only on CL_CGM_. When a participant showed at least one positive measure at baseline, the number of positive measures tended to increase at follow-up, with only one individual (of 63) showing a decrease when over four measures were positive at baseline (Figure S9).

Figure 3C shows ARC (CL/year) for all measures across baseline ConcN, Disc and ConcP groups. For all longitudinal measures, the mean ARC in the baseline ConcN, Disc and ConcP groups was between 0.1–1.4, 3.2–4.6 and 3.0–4.7 CL/year, respectively. All measures showed significant group differences using Welch’s ANOVA (all *p* < 0.001 after FDR correction). ConcN showed significantly lower ARC compared to both Disc and ConcP for all measures (Games-Howell post-hoc, all *p* < 0.05 FDR-adjusted, Table S5). We did not observe any significant difference in ARC between Disc and ConcP groups, with mean differences close to zero across measures (-0.2–0.2 CL/yr, Table S5). Figure 3D shows the proportion of reliable accumulators (rate of change above the reliable accumulation cut-point for each method) across each baseline concordance group. The proportion of reliable accumulators was low in the baseline ConcN group (n = 10–20, 4.2–8.5% across measures) and higher in the Disc (n = 19–32, 34.5–58.2%) and ConcP (n = 13–21, 39.4–63.6%) groups. Fisher’s exact tests found significantly higher odds of reliable accumulator classification for Disc and ConcP groups compared to ConcN with no significant differences between Disc and ConcP groups for any measure (Table S6).

### 3.5 PET discordance and CSF Aβ42/Aβ40

There were 134 individuals with CSF Aβ42/Aβ40 available at phase 2 (contemporaneous with follow-up PET scan), where 20 (14.9%) and 19 (14.2%) individuals were classified as likely positive and positive with FDA published cut-points, respectively. A subset of these individuals (n = 120) had baseline PET, 16 (13.3%) and 18 (15.0%) of whom were classified as likely positive and positive on CSF. CSF was available in 113 individuals that also had follow-up PET, with 16 (14.2%) and 16 (14.2%) individuals classified as CSF likely positive and positive.

In the CSF subset at baseline (n = 120), 83 (69.2%) were ConcN, 25 (20.8%) were Disc and 12 (10%) were ConcP. At follow-up (n = 113), 75 (66.4%) were ConcN, 20 (17.7%) were Disc and 18 (15.9%) were ConcP. Figure 4A shows CSF Aβ42/Aβ40 by PET concordance group. We found a significant difference in CSF Aβ42/Aβ40 between baseline concordance groups (Welch’s ANOVA, *p* < 0.001) with Games-Howell post-hoc tests showing significantly lower CSF Aβ42/Aβ40 in Disc and ConcP groups compared to ConcN (both *p* < 0.001) and in ConcP compared to Disc (*p* = 0.007). Individuals who were rated Disc at baseline or follow-up were likely to have abnormal CSF Aβ42/Aβ40 and all individuals that were ConcP at baseline or follow-up were CSF Aβ42/Aβ40 abnormal (ratio < 0.073). The heatmap presented in Figure 4B visualises the PET status for all measures and each individual who was categorised as discordant at either baseline or follow-up, ordered by CSF level. In the normal CSF range (≥ 0.073), the two discordant cases at baseline with only one positive measure (CGM_PVC) who are subsequently ConcN are likely false positives, but discordant cases with increasing number of positive measures at follow-up are more likely to be true positives – particularly at lower, more abnormal CSF levels.

**Figure 4.**
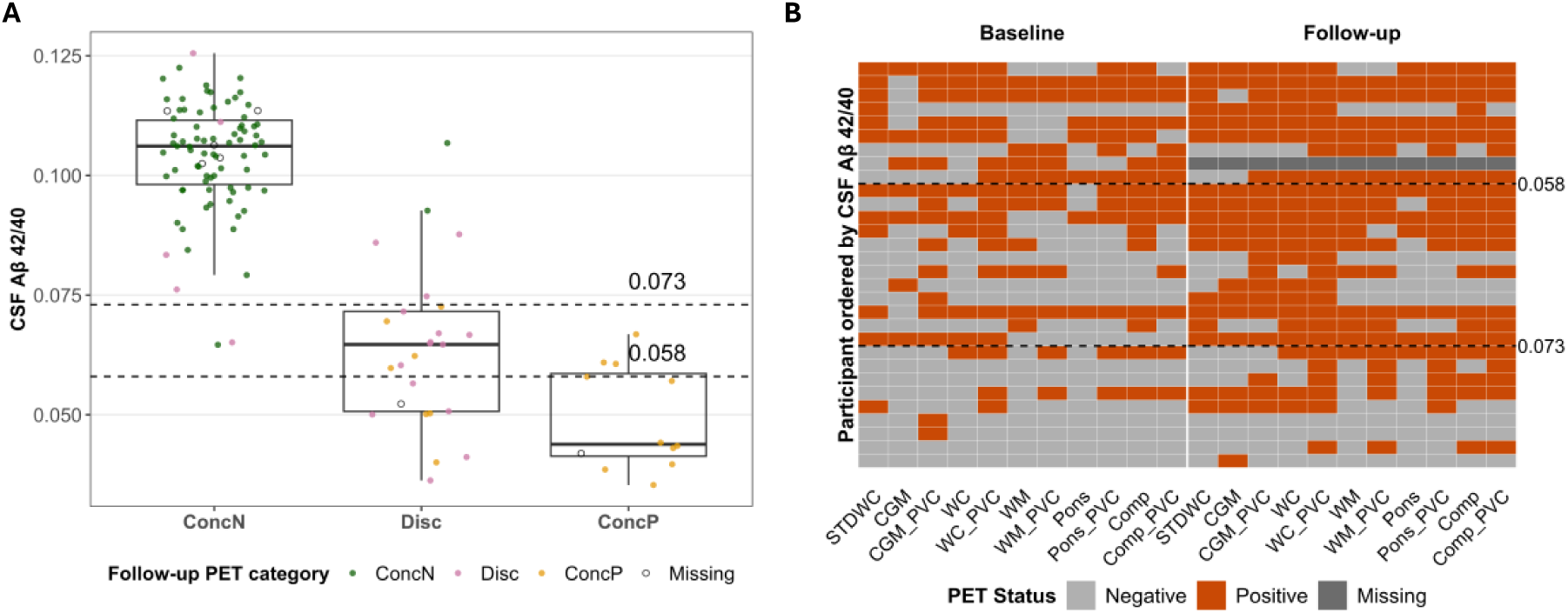
PET discordance and CSF Aβ42/40. **A**: CSF Aβ42/40 at follow-up by baseline PET concordance groups. Points are coloured according to their PET concordance category at ∼2.5-year follow-up, indicating whether they later transitioned or remained in the same category. **B**: The colour of each cell represents PET status at baseline (left) or follow-up (right) across measures in individuals with CSF that were PET discordant at either baseline or follow-up. Dashed lines represent the published FDA cut-points for the CSF Aβ42/40 Fujirebio Lumipulse ratio (positive ≤0.058, likely positive 0.059–0.072, negative ≥ 0.073).

## 4 Discussion

We evaluated how PET processing methodology influences Aβ quantification and its uncertainty, within a predominantly cognitively unimpaired community cohort. Different reference regions and the implementation of PVC produced meaningful differences in Centiloid cut-points, positivity rates and cut-point stability. Discordant Aβ status across methods (18%) was more common than concordant positivity (12%) and occurred primarily within the 11–35 CL range. PET and CSF profiles at follow-up roughly two and a half years later suggested that most discordant individuals represent early Aβ accumulation.

### 4.1 Cut-point estimates and their uncertainty

A key aim of this study was to determine how Aβ PET cut-points and their stability vary across commonly used processing methods. Baseline Centiloid distributions were effectively bi-modal for all measures, which resulted in GMM-derived cut-points between 10 and 23 CL. This range lies within the lower end of the previously described “evolving amyloid” zone, consistent with the use of a data-driven GMM approach in a largely cognitively unimpaired cohort[11,29]. Positivity estimates ranged from 16–25%, highlighting substantial methodological variability. WM and pons reference regions produced more conservative positivity estimates than cerebellar reference regions. The highest positivity estimates were consistent with a recent meta-analysis reporting 24% (95% CI 21–28%) Aβ positivity at age 70 in cognitively unimpaired individuals[30]. Application of PVC increased positivity rates for CGM and WC reference regions, likely reflecting correction for white matter spill-in to cortical target regions.

Processing methodology also influenced estimates of reliable Aβ accumulation. Reliable accumulation cut-points ranged from 3.5–6 CL/year for CGM, WC (with and without PVC) and the composite reference region without PVC. In contrast, WM, pons (with and without PVC) and the composite reference region with PVC resulted in better fits with a unimodal Gaussian distribution, precluding GMM-derived accumulation thresholds. Our GMM-derived threshold for standard Centiloids (3.9 CL/year) was slightly higher than the previous Insight 46 estimate (3.2 CL/year) but remained within its 95% confidence interval, likely reflecting updated PET reconstruction[13]. The lower threshold observed in AMYPAD-PNHS (2.2 CL/year) may reflect differences in cohort characteristics or image acquisition[13]. PVC substantially improved the stability of accumulation estimates for the CGM reference region but provided little benefit for other reference regions. Together, these findings demonstrate that reference region selection, PVC and the interaction between them substantially influence estimates of reliable Aβ accumulation.

### 4.2 Discordance is suggestive of early amyloidosis

Although most individuals (82%) were classified consistently across PET processing methods, discordance (18%) was slightly more common than concordant positivity (12%) in this largely cognitively unimpaired cohort. Discordance occurred predominantly between 11 and 35 CL using the standard Centiloid method with individuals below or above those values when using the standard approach likely to be rated negative or positive regardless of methodology used. This range corresponds closely to the intermediate Centiloid range used in preclinical prevention trials[3], the range with greatest inter-reader disagreement[5,31], and the location of most GMM derived cut-points in a recent large meta-analysis of CL values across studies including Insight 46[11].

CSF and PET measurements at follow-up helped resolve the biological significance of discordance. Discordant individuals transitioned to concordant positivity approximately 2.7 times more frequently than they reverted to concordant negativity, showed progressively higher APOE ε4 frequency across ConcN, Disc and ConcP groups, with CSF Aβ 42/40 ratios at follow-up and longitudinal changes in PET resembling concordant positives. Furthermore, all reversions (Disc->ConcN or ConcP->Disc) were driven by disagreement of a single PET measure while the remaining measures agreed. Together, these findings suggest that most discordance reflects transitional Aβ accumulation rather than methodological error, although this does not preclude isolated discordance that may still arise from technical artefact.

These findings suggest that combining multiple PET quantification approaches may improve sensitivity to early Aβ accumulation in research settings, although this is unlikely to be practical clinically. The biological and technical factors underlying discordance warrant further investigation.

### 4.3 Implications for defining Aβ abnormality in research and trials

The Centiloid framework was developed as a post-hoc transformation allowing research groups to retain their preferred processing methods while expressing results on a common scale[4,32]. Our findings demonstrate that substantial differences remain between processing methods, even when these use many of the same parameters (e.g. brain parcellation and co-registration methodologies) and radiotracer, scanner and reconstruction are held constant.

The optimal processing strategy depends on study design. Methods that perform well in single-site studies with standardised acquisition may not generalise to large multicentre studies using multiple tracers and scanners[33,34]. Our findings suggest that well-implemented PVC can improve sensitivity when cerebellar reference regions are used, although in multicentre datasets with variable scanner resolution or limited MRI quality, PVC may instead increase measurement variability. Previous studies have suggested that reference regions containing subcortical white matter provide sensitive longitudinal measurements[33,35–37]. The relatively poor longitudinal performance of WM in this cohort was therefore unexpected and may reflect unresolved biological or technical factors affecting non-specific white matter uptake. In contrast, the good performance of the composite reference region without PVC agrees with previous recommendations for multicentre datasets such as ADNI, where cerebellar signal alone may be less reliable because of variation in scanner characteristics[33,38].

### 4.4 Strengths and limitations

Strengths of this study include evaluation of all commonly used PET reference regions together with PVC, and the availability of CSF and longitudinal PET biomarkers that enabled biological interpretation of discordant cases. The relatively large, community-based Insight 46 sample is likely more representative of secondary prevention populations than clinic-based cohorts, while its narrow age range minimises confounding by age[6].

Several limitations should also be acknowledged. The cohort is entirely White European and does not fully represent the contemporary UK population of 70-year-olds due to demographic changes since the mid-1940s, potentially limiting generalisability. Participants also tend to have higher educational attainment and better health than the wider birth cohort[39]. Finally, the use of a single scanner and radiotracer may underestimate methodological variability compared with multicentre studies incorporating multiple scanners and tracers. Future work should examine whether these findings generalise to more diverse samples, such as the 1958 British birth cohort, and imaging protocols, with a further wave of data collection currently underway using [^18^F]florbetaben in a different group of individuals from the 1946 British birth cohort.

### 4.5 Conclusion

Processing methodology introduces substantial uncertainty into amyloid PET quantification, particularly within the 11–35 Centiloid range where processing methods frequently disagree. Longitudinal PET and CSF data suggest that most discordant cases represent early Aβ accumulation rather than methodological error. These findings highlight the importance of accounting for methodological uncertainty when defining Aβ positivity in secondary prevention trials and other studies of preclinical Alzheimer’s disease.

## Data Availability

All data produced in the present study are available upon reasonable request to the authors.

https://condor.ucl.ac.uk/Condor/

## CRediT authorship contribution statement

**William Coath:** Writing – review & editing, Writing – original draft, Visualization, Software, Methodology, Investigation, Formal analysis, Data curation, Conceptualization. **Ariane Bollack:** Writing – review & editing, Software, Conceptualization. **Catherine J Scott:** Writing – review & editing, Data Curation, Software, Conceptualization. **Ashvini Keshavan:** Writing – review & editing, Investigation, Data Curation. **Ian B Malone:** Writing – review & editing, Software, Data Curation. **Heidi Murray-Smith:** Writing – review & editing, Project administration, Data Curation, Resources, Investigation. **Pawel J Markiewicz:** Writing – review & editing, Software, Data Curation. **Kjell Erlandsson:** Writing – review & editing, Software. **Benjamin A Thomas:** Writing – review & editing, Software. **Frederik Barkhof:** Writing – review & editing, Resources, Methodology. **John C Dickson:** Writing – review & editing, Resources, Methodology, Funding acquisition. **Michael Schöll:** Writing – review & editing, Supervision, Conceptualization. **Jonathan M Schott:** Writing – review & editing, Writing – original draft, Supervision, Resources, Methodology, Funding acquisition, Conceptualization. **David M Cash:** Writing – review & editing, Writing – original draft, Supervision, Resources, Methodology, Funding acquisition, Conceptualization.

## Acknowledgements

J.M.S. is a National Institute for Health Research (NIHR) Senior Investigator and acknowledges the support of the NIHR University College London Hospitals Biomedical Research Centre and the UCL Centre of Research Excellence, an initiative funded by British Heart Foundation (RE/24/130013). He has grant funding from Alzheimer’s Research UK, Brain Research UK, Weston Brain Institute, Medical Research Council, British Heart Foundation, Wolfson Foundation, UK Dementia Research Institute, and Alzheimer’s Association. This work uses data provided by study participants or patients and collected through clinical studies or as part of their care and support. Insight 46 is funded by grants from Alzheimer’s Research UK (ARUK-PG2014-1946 and ARUK-PG2017-1946), Alzheimer’s Association (SG-666374-UK BIRTH COHORT), the Medical Research Council Dementias Platform UK (CSUB19166), The Wolfson Foundation (PR/ylr/18575), The Medical Research Council (MC_UU_10019/1 and MC_UU_10019/3) and Brain Research Trust (UCC14191). Florbetapir amyloid tracer was provided in kind by AVID Radiopharmaceuticals (a wholly owned subsidiary of Eli Lilly), who had no part in the design of the study. The funders of the study had no role in study design, data collection, analysis, interpretation, report writing or in the decision to submit the article for publication.

D.M.C. is supported by an Alzheimer’s Society Dementia Research Leaders Fellowship (AS-DRL-23-005), Alzheimer’s Association (SG-666374-UK BIRTH COHORT), the National Institute for Health and Care Research University College London Hospitals Biomedical Research Centre, and the UK Dementia Research Institute, which receives its funding from DRI Ltd, funded by the UK Medical Research Council, Alzheimer’s Society, and Alzheimer’s Research UK.

## Declaration of generative AI and AI-assisted technologies in the manuscript preparation process

During the preparation of this work, the author(s) used ChatGPT for proof reading and editing of text for clarity and conciseness. The author(s) reviewed and edited the output as needed and take full responsibility for the content of the published article.

