## Supplemental information for "Amyloid-PET pipeline choice influences classification of preclinical Alzheimer’s disease"

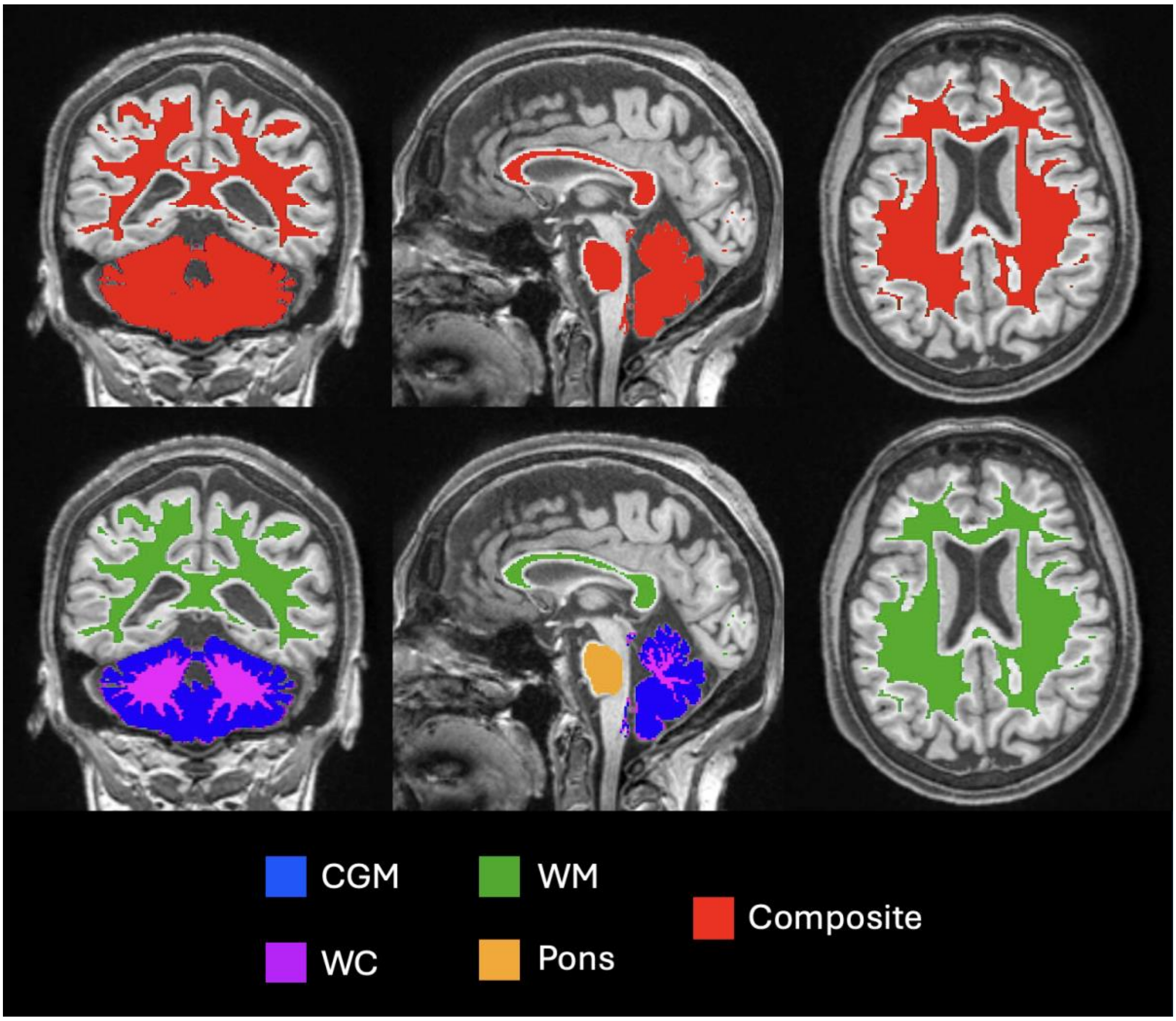


*Figure S1. Example of GIF based reference regions overlaid on T1w MRI. Orientation: The left of the image is on the participant's right. Abbreviations: CGM = cerebellar grey matter; WC = Whole cerebellum (encompasses the CGM region and cerebellar WM); WM = Eroded subcortical white matter; GIF = Geodesic information flows parcellation; PET = positron emission tomography.*

*Table S1. Centiloid (CL) conversion equations for each measure. CL = SUVR*slope + intercept. The equation for conversion for florbetapir SUVRs using the standard approach (STDWC) is published in Navitsky et al. (2018).*

| Measure | Slope | Intercept |
|---|---|---|
| STDWC | 175.0 | -182.0 |
| CGM | 159.1 | -186.3 |
| CGM_PVC | 79.3 | -93.3 |
| WC | 194.0 | -194.7 |
| WC_PVC | 106.9 | -98.4 |
| WM | 431.1 | -237.4 |
| WM_PVC | 221.2 | -126.6 |
| Pons | 338.0 | -196.7 |
| Pons_PVC | 191.1 | -115.4 |
| Comp | 282.8 | -215.6 |
| Comp_PVC | 175.7 | -122.1 |

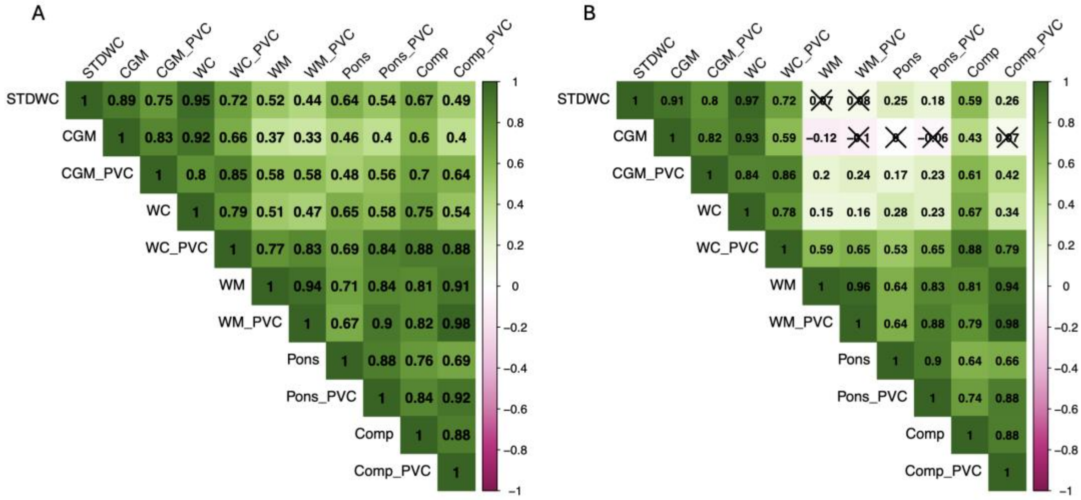


*Figure S2. Non-parametric Spearman's rank correlation between methods at baseline (A) and in annual rate of change (B). Squares marked with cross were p > .05 uncorrected for multiple comparisons.*

*Table S2. Bayesian information criterion (BIC) differences between model types at baseline. BIC difference >6 from the optimal model (BEST) is considered strong evidence and highlighted in red (Kass & Raftery, 1995). Model type codes: 1 = unimodal; 2E = Two Gaussian with equal variance terms; 2V = Two Gaussian with unequal variance terms; 3E = Three Gaussian with equal variance terms; 3V = Three Gaussian with unequal variance terms.*

| **Measure** | **Model type** | | | | |
|---|---|---|---|---|---|
| | **1** | **2E** | **2V** | **3E** | **3V** |
| STDWC | 246.5 | 38.8 | **BEST** | 51.0 | 9.5 |
| CGM | 235.1 | 43.4 | **BEST** | 55.5 | 5.6 |
| CGM_PVC | 402.3 | 130.0 | 0.7 | 142.2 | **BEST** |
| WC | 323.3 | 80.4 | **BEST** | 92.6 | 6.3 |
| WC_PVC | 485.0 | 171.0 | 16.2 | 183.1 | **BEST** |
| WM | 188.3 | 35.0 | **BEST** | 47.2 | 11.9 |
| WM_PVC | 246.9 | 53.2 | **BEST** | 65.4 | 7.8 |
| Pons | 228.4 | 21.6 | **BEST** | 33.8 | 14.7 |
| Pons_PVC | 322.2 | 64.0 | 7.2 | 76.2 | **BEST** |
| Comp | 360.1 | 104.2 | **BEST** | 116.3 | 3.9 |
| Comp_PVC | 311.1 | 65.5 | **BEST** | 77.7 | 3.0 |

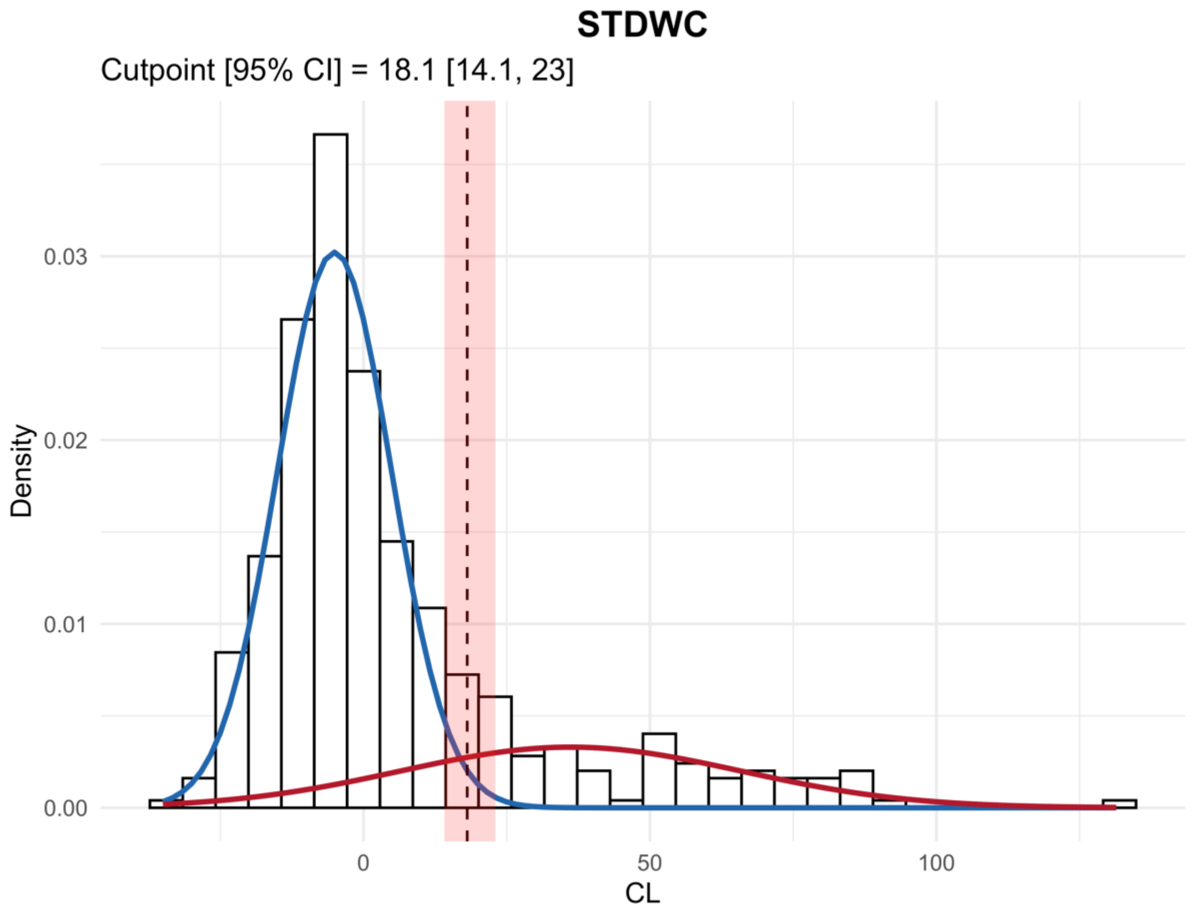

*Figure S3. Baseline Centiloid measured with the standard processing (STDWC).*

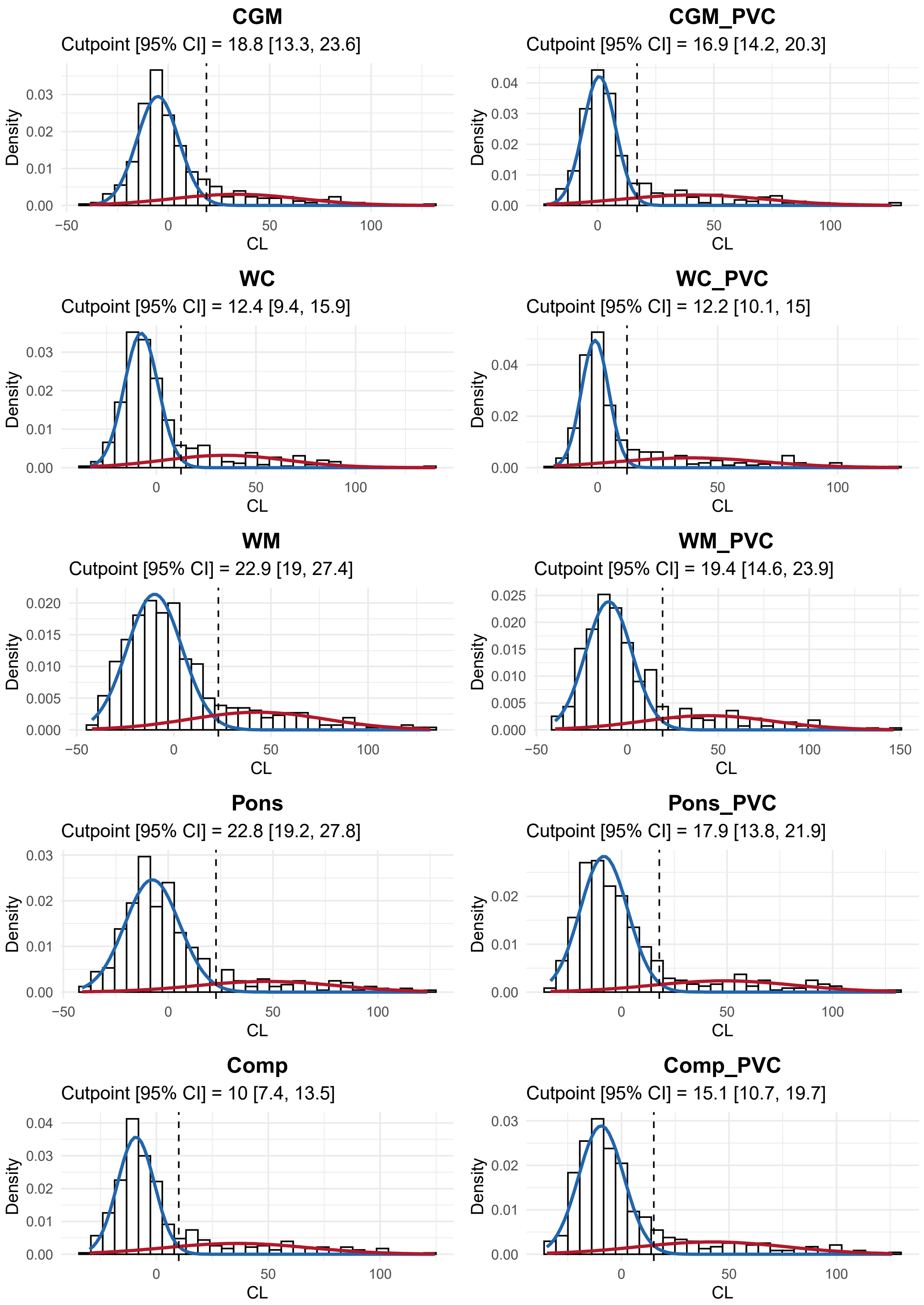


*Figure S4. Non-standard measure GMM fits at baseline (n = 433).*

*Table S3. Centiloid cutpoints and rates.*

| Measure | Positivity cut-point<br>CL [95% CI] | Baseline positivity rate<br>% [95% CI] | Reliable accumulator cut-point<br>CL/yr [95% CI] | Reliable accumulator rate<br>% [95% CI] |
|---|---|---|---|---|
| STDWC | 18.1 [14.1, 23.0] | 19.4 [16.2, 21.7] | 3.9 [2.4, 4.8] | 19.8 [15.7, 31.2] |
| CGM | 18.8 [13.3, 23.6] | 17.1 [14.3, 21.2] | 6.2 [2.4, 9.1] | 16.0 [5.9, 43.8] |
| CGM_PVC | 16.9 [14.2, 20.3] | 22.2 [20.3, 24.7] | 4.9 [3.5, 6.4] | 13.0 [8.3, 19.8] |
| WC | 12.4 [9.4, 15.9] | 19.6 [18.0, 22.6] | 4.0 [2.3, 5.2] | 22.2 [16.4, 37.0] |
| WC_PVC | 12.2 [10.1, 15.0] | 25.2 [23.6, 27.3] | 3.5 [2.1, 4.7] | 21.3 [14.5, 33.6] |
| WM | 22.9 [19.0, 27.4] | 18.2 [16.9, 19.9] | NA | NA |
| WM_PVC | 19.4 [14.6, 23.9] | 18.7 [17.1, 21.7] | NA | NA |
| Pons | 22.8 [19.2, 27.8] | 15.9 [14.1, 17.1] | NA | NA |
| Pons_PVC | 17.9 [13.8, 21.9] | 17.1 [15.0, 19.6] | NA | NA |
| Comp | 10.0 [7.4, 13.5] | 22.2 [21.0, 24.0] | 3.9 [0.9, 5.9] | 21.0 [10.2, 53.4] |
| Comp_PVC | 15.1 [10.7, 19.7] | 19.9 [17.3, 22.9] | NA | NA |

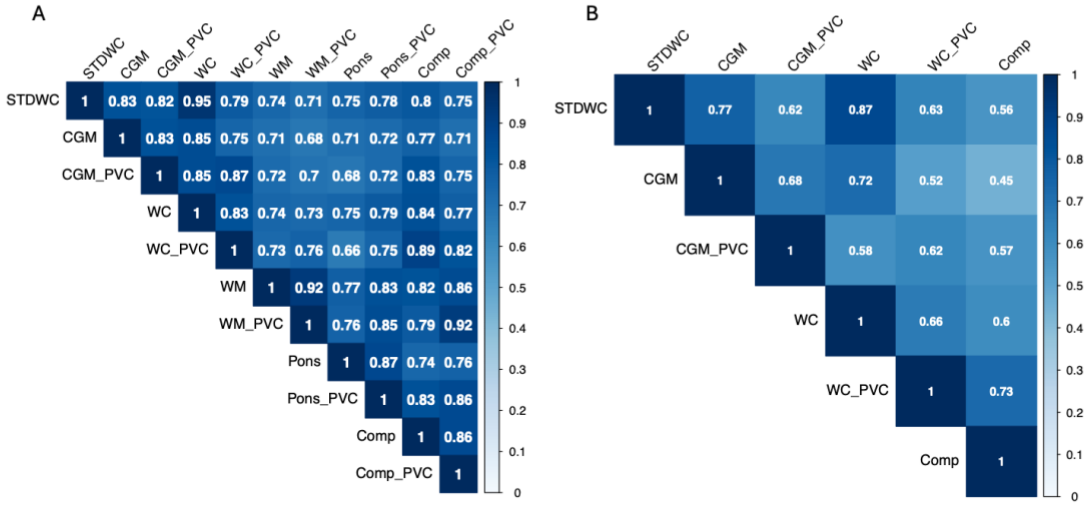


*Figure S5. Agreement (kappa) between methods in positivity (A) and accumulator (B) status.*

*Table S4. Bayesian information criterion (BIC) differences between model types with annual rate of change. BIC difference >6 is considered strong evidence (Kass & Raftery, 1995). Model type codes: 1 = unimodal; 2E = Two Gaussian with equal variance terms; 2V = Two Gaussian with unequal variance terms; 3E = Three Gaussian with equal variance terms; 3V = Three Gaussian with unequal variance terms. *Methods which favoured the single Gaussian model were excluded from the longitudinal analysis.*

| **Measure** | **Model Type** | | | | |
|---|---|---|---|---|---|
| | **1** | **2E** | **2V** | **3E** | **3V** |
| STDWC | 37.4 | 27.7 | **BEST** | 39.3 | 14.0 |
| CGM | 7.7 | 7.4 | **BEST** | 18.9 | 14.8 |
| CGM_PVC | 20.7 | 7.4 | **BEST** | 19.0 | 17.3 |
| WC | 40.0 | 21.2 | **BEST** | 32.9 | 15.4 |
| WC_PVC | 33.9 | 6.3 | **BEST** | 18.0 | 16.7 |
| WM* | **BEST** | 11.8 | 11.6 | 17.3 | 29.4 |
| WM_PVC* | **BEST** | 6.6 | 11.1 | 18.1 | 28.6 |
| Pons* | **BEST** | 6.8 | 12.2 | 18.7 | 29.2 |
| Pons_PVC* | **BEST** | 4.9 | 10.7 | 16.5 | 28.2 |
| Comp | 13.8 | **BEST** | 1.0 | 11.6 | 12.0 |
| Comp_PVC* | **BEST** | 4.4 | 8.8 | 16.1 | 26.0 |

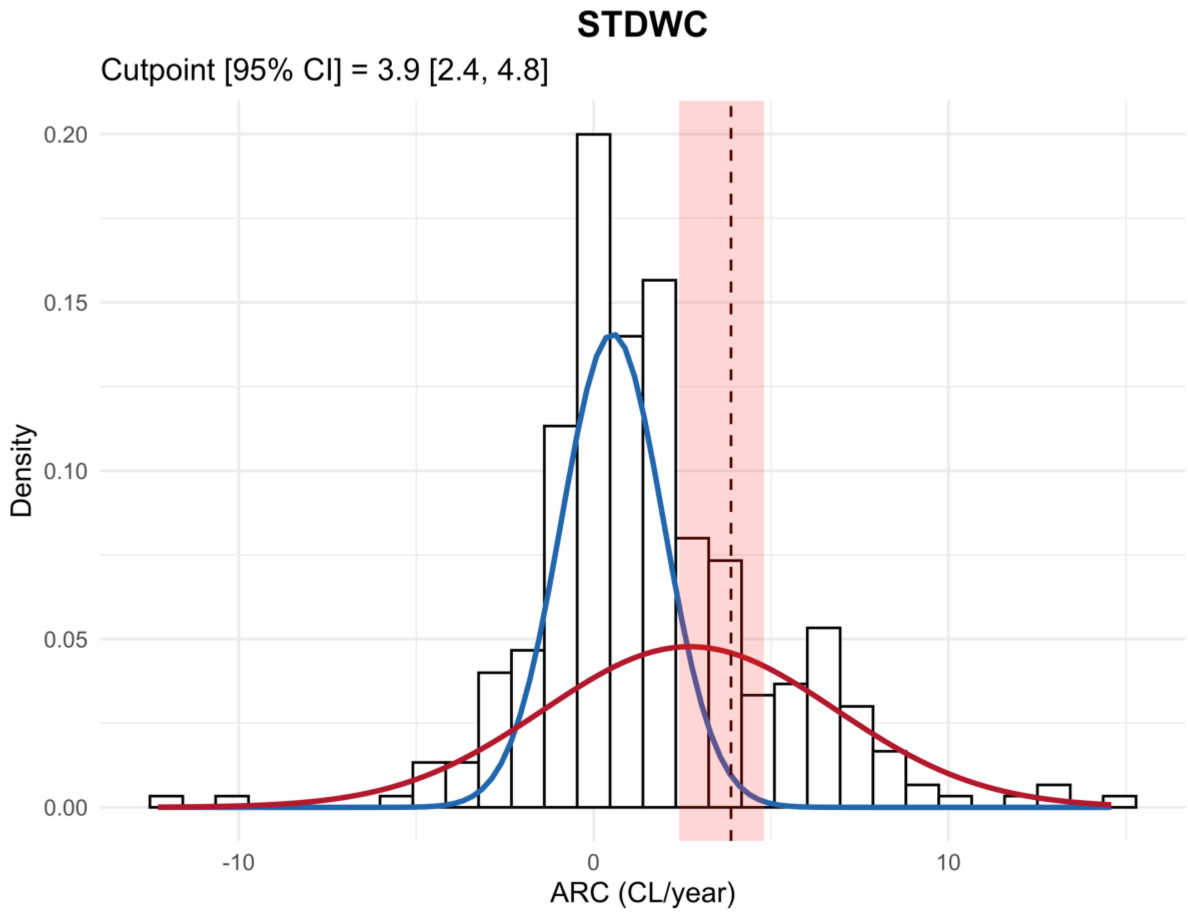


*Figure S6. Annual rate of change measured with the standard Centiloid method (STDWC).*

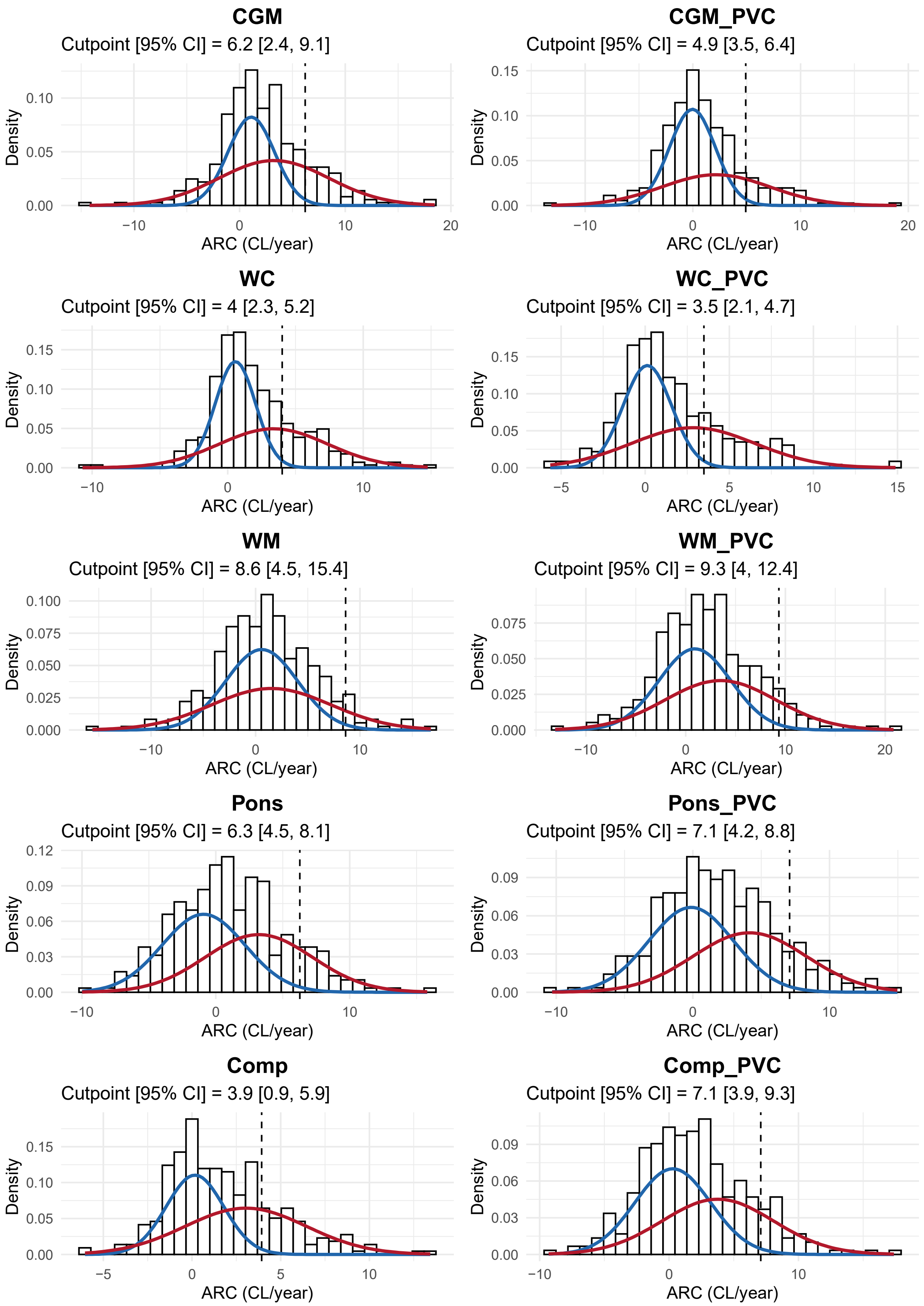


*Figure S7. Non-standard measure GMM fits for annual rate of change (ARC) (n = 324).*

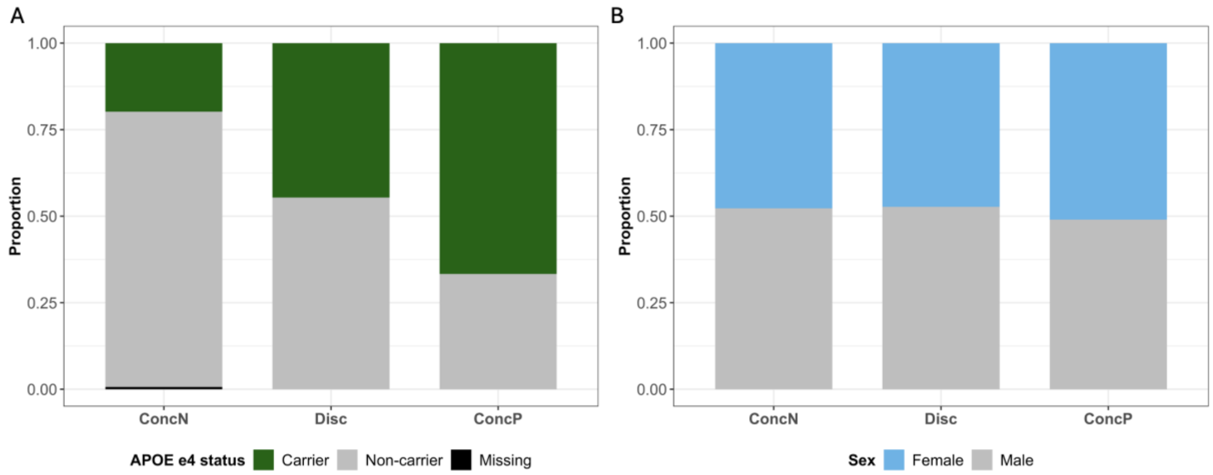


*Figure S8. Proportion of APOE e4 carriers (A) and females (B) in each baseline concordance category.*

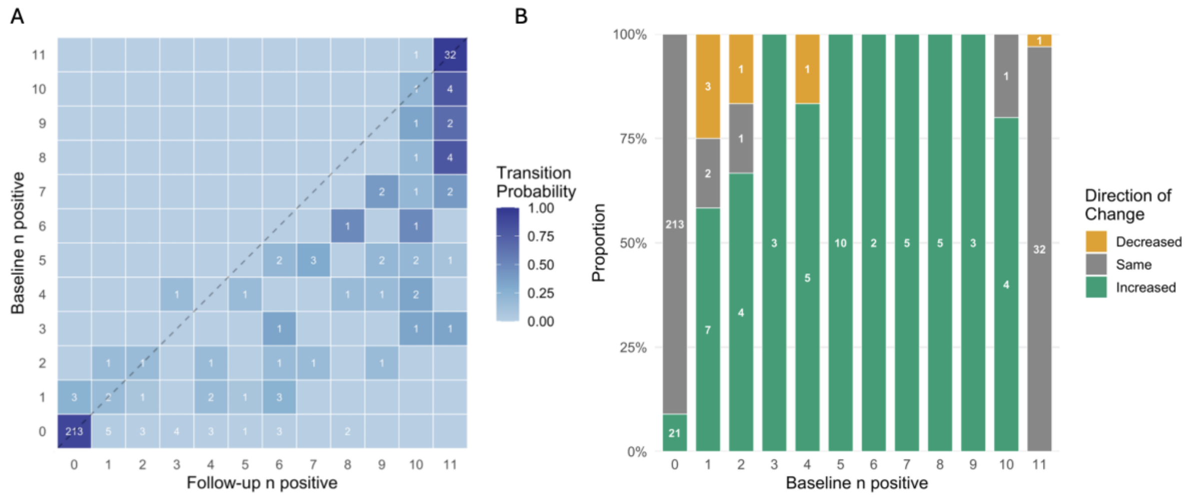


*Figure S9. Number of positive measures at baseline and follow-up in individuals with longitudinal data (n = 324). A: Heatmap showing pattern of transitions between measures with deeper blue indicating higher probability (calculated row-wise) and numbers indicated on tiles. B: The proportion of individuals where the number of positives decreased, increased and stayed the same between timepoints. Decreases were rare when >2 measures were positive at baseline, with only a single case when >4 measures were positive.*

*Table S5. Differences in annual rate of change in amyloid between baseline concordance groups. Games-Howell post-hoc tests following significant Welch's ANOVA *p < 0.05 and **p < 0.001 FDR adjusted.*

| Measure | Comparison | CL/yr difference estimate [95% CI] | *p* value |
|---|---|---|---|
| STDWC | **ConcN vs Disc** | **3.3 [2.1, 4.4]** | **<0.001**** |
| | **ConcN vs ConcP** | **3.4 [1.0, 5.7]** | **0.004*** |
| | Disc vs ConcP | 0.1 [-2.5, 2.7] | 0.995 |
| CGM | **ConcN vs Disc** | **3.1 [1.6, 4.7]** | **<0.001**** |
| | **ConcN vs ConcP** | **3.3 [0.3, 6.3]** | **0.030*** |
| | Disc vs ConcP | 0.2 [-3.1 , 3.5] | 0.992 |
| CGM_PVC | **ConcN vs Disc** | **3.1 [1.6, 4.6]** | **<0.001**** |
| | **ConcN vs ConcP** | **2.9 [0.2, 5.7]** | **0.037*** |
| | Disc vs ConcP | -0.2 [-3.2, 2.9] | 0.992 |
| WC | **ConcN vs Disc** | **3.6 [2.4, 4.7]** | **<0.001**** |
| | **ConcN vs ConcP** | **3.6 [1.3, 6.0]** | **0.002*** |
| | Disc vs ConcP | 0.1 [-2.5, 2.6] | 0.997 |
| WC_PVC | **ConcN vs Disc** | **3.8 [2.6, 4.9]** | **<0.001**** |
| | **ConcN vs ConcP** | **3.7 [1.9, 5.6]** | **<0.001**** |
| | Disc vs ConcP | 0.0 [-2.1 , 2.1] | 0.999 |
| Comp | **ConcN vs Disc** | **3.9 [2.8, 5.0]** | **<0.001**** |
| | **ConcN vs ConcP** | **4.0 [2.4, 5.5]** | **<0.001**** |
| | Disc vs ConcP | 0.1 [-1.7, 1.9] | 0.992 |

*Table S6. The association between reliable accumulator status and baseline concordance group. Fisher's exact tests *p < 0.05 and **p < 0.001 FDR adjusted.*

| Measure | Comparison | Odds ratio [95% CI] | *p* value |
|---|---|---|---|
| STDWC | **ConcN vs Disc** | **17.2 [7.8 , 39.8]** | **<0.001**** |
| | **ConcN vs ConcP** | **23.5 [9.1, 64.1]** | **<0.001**** |
| | Disc vs ConcP | 1.4 [0.5, 3.6] | 0.579 |
| CGM | **ConcN vs Disc** | **7.3 [3.3, 16.6]** | **<0.001**** |
| | **ConcN vs ConcP** | **12.6 [5.0, 32.5]** | **<0.001**** |
| | Disc vs ConcP | 1.7 [0.7, 4.6] | 0.368 |
| CGM_PVC | **ConcN vs Disc** | **11.1 [4.5, 28.8]** | **<0.001**** |
| | **ConcN vs ConcP** | **14.3 [5.1, 41.7]** | **<0.001**** |
| | Disc vs ConcP | 1.3 [0.5, 3.4] | 0.688 |
| WC | **ConcN vs Disc** | **14.3 [6.8, 31.1]** | **<0.001**** |
| | **ConcN vs ConcP** | **19.4 [7.8, 50.8]** | **<0.001**** |
| | Disc vs ConcP | 1.4 [0.5, 3.7] | 0.579 |
| WC_PVC | **ConcN vs Disc** | **18.4 [8.5, 41.6]** | **<0.001**** |

| | | | |
|---|---|---|---|
| | **ConcN vs ConcP** | **20.5 [8.1, 54.4]** | **<0.001**** |
| | Disc vs ConcP | 1.1 [0.4, 3.0] | 0.828 |
| Comp | **ConcN vs Disc** | **18.5 [8.6, 41.6]** | **<0.001**** |
| | **ConcN vs ConcP** | **13.3 [5.3, 34.1]** | **<0.001**** |
| | Disc vs ConcP | 0.7 [0.3, 1.9] | 0.579 |
